# Dual-phase vessel wall MRI deep learning for identifying composite unstable intracranial aneurysm phenotypes: a multicenter study

**DOI:** 10.64898/2026.08.13.26360349

**Authors:** Wenqing Yuan, Zihang Wang, Qingyu Wu, Xiaoyu He, Jiaquan Tan, Xin Wei, Runxin Li, Ying Yin, Dongdong Wang, Guangxian Wang, Ting Chen

## Abstract

**Objectives:** To develop and externally validate a wall-focused deep learning framework for identifying composite unstable intracranial aneurysm phenotypes on dual-phase high-resolution vessel wall imaging (HR-VWI), and to visualize model attention on the aneurysm wall surface.

**Methods:** This retrospective multicenter study included patients with intracranial aneurysms who underwent both non-contrast and contrast-enhanced HR-VWI. Center 1 was used for model development and patient-level five-fold out-of-fold assessment, whereas Centers 2 and 3 served as independent external validation cohorts. For each aneurysm, dual-phase local wall patches and larger spatial context patches were generated. The Wall-Constrained Encoding Network (WCE-Net) extracted mask-constrained local wall features, and a transfer-learning U-Net with Nested Transformers (UNesT) branch extracted spatial context information. Branch outputs were fused by logit-level stacking. Model performance was evaluated using discrimination, calibration, and decision curve analysis. Three-dimensional gradient-weighted class activation mapping (Grad-CAM) responses were projected onto the reconstructed aneurysm wall surface and compared with HR-VWI surface signal intensity.

**Results:** A total of 629 patients with 773 aneurysms were included. The final fusion model achieved areas under the receiver operating characteristic curves (AUCs) of 0.908, 0.857, and 0.855 in Center 1, external Center 2, and external Center 3, respectively. Corresponding Brier scores were 0.119, 0.153, and 0.150. Surface Grad-CAM showed partial spatial overlap between model-attention hotspots and high-signal HR-VWI regions.

**Conclusions:** Dual-phase wall-focused local-context fusion showed feasibility for identifying composite unstable intracranial aneurysm phenotypes across centers. Surface Grad-CAM provided anatomically referenced visualization of model attention.

**Key Points:** 

**Question:** Can dual-phase high-resolution vessel wall imaging deep learning consistently identify composite unstable intracranial aneurysm phenotypes across internal and independent external cohorts?

**Findings:** The fusion model showed consistent discrimination across internal and independent external cohorts and enabled anatomically referenced visualization of model attention.

**Critical Relevance Statement:** External validation supports dual-phase wall-focused deep learning for cross-center vessel wall MRI assessment of composite unstable aneurysm phenotypes and anatomically referenced visualization of model attention.

## 1. Introduction

Rupture of intracranial aneurysms can cause subarachnoid hemorrhage and is associated with high mortality and disability [1]. Identifying unstable phenotypes among unruptured aneurysms is essential for optimizing treatment decisions [2,3].

High-resolution vessel wall imaging (HR-VWI) can directly depict aneurysm wall structure and enhancement characteristics, providing an important imaging basis for assessing aneurysm instability [4]. Non-contrast and contrast-enhanced sequences may provide complementary information by reflecting baseline wall signal and enhancement-related features, respectively. However, visual assessment is influenced by reader experience, and a single wall-enhancement sign may not sufficiently characterize complex and heterogeneous aneurysm wall phenotypes [5–8]. Therefore, stable extraction of imaging phenotypes related to aneurysm instability from vessel wall imaging remains an important issue for improving risk assessment.

Radiomics and deep learning methods have been applied to intracranial aneurysm detection, segmentation, and risk assessment [9–14], but existing approaches often rely on predefined features or general-purpose three-dimensional encoders [12–14]. Aneurysm walls are thin, small in volume, irregular in shape, and susceptible to adjacent background signals; aneurysm instability may also be influenced by surrounding anatomical context [8]. An ideal model should therefore integrate local aneurysm wall features and broader spatial context while providing spatial explanations referenced to the aneurysm wall.

In this study, we developed a WCE-Net-based wall-focused deep learning framework for identifying composite unstable intracranial aneurysm phenotypes on dual-phase HR-VWI. The framework combines a WCE-Net local wall encoder with a transfer-learning UNesT context encoder to integrate local aneurysm wall phenotypes and broader spatial context, and was evaluated using internal out-of-fold assessment and two independent external validation cohorts [15]. To improve anatomical visualization, three-dimensional Grad-CAM [16] responses were projected onto the aneurysm wall surface and spatially compared with HR-VWI surface signal intensity, allowing model attention to be inspected in relation to wall-surface imaging patterns.

## 2. Methods

### 2.1 Study Design and Patient Cohorts

This multicenter retrospective study was approved by the Ethics Committee of the Second Affiliated Hospital of Chongqing Medical University (approval/reference No. 194) and was conducted in accordance with the Declaration of Helsinki and its subsequent amendments. The requirement for written informed consent was waived by the ethics committee due to the retrospective nature of the study and the use of anonymized clinical and imaging data. Patients with intracranial aneurysms from three centers between January 2022 and January 2026 were retrospectively screened; all patients underwent HR-VWI before and after contrast enhancement. January 2026 represented the final imaging acquisition date rather than the endpoint of clinical follow-up. Figure 1 shows the patient enrollment flowchart, and Figure 2 shows the overall study workflow.

**Figure 1.**
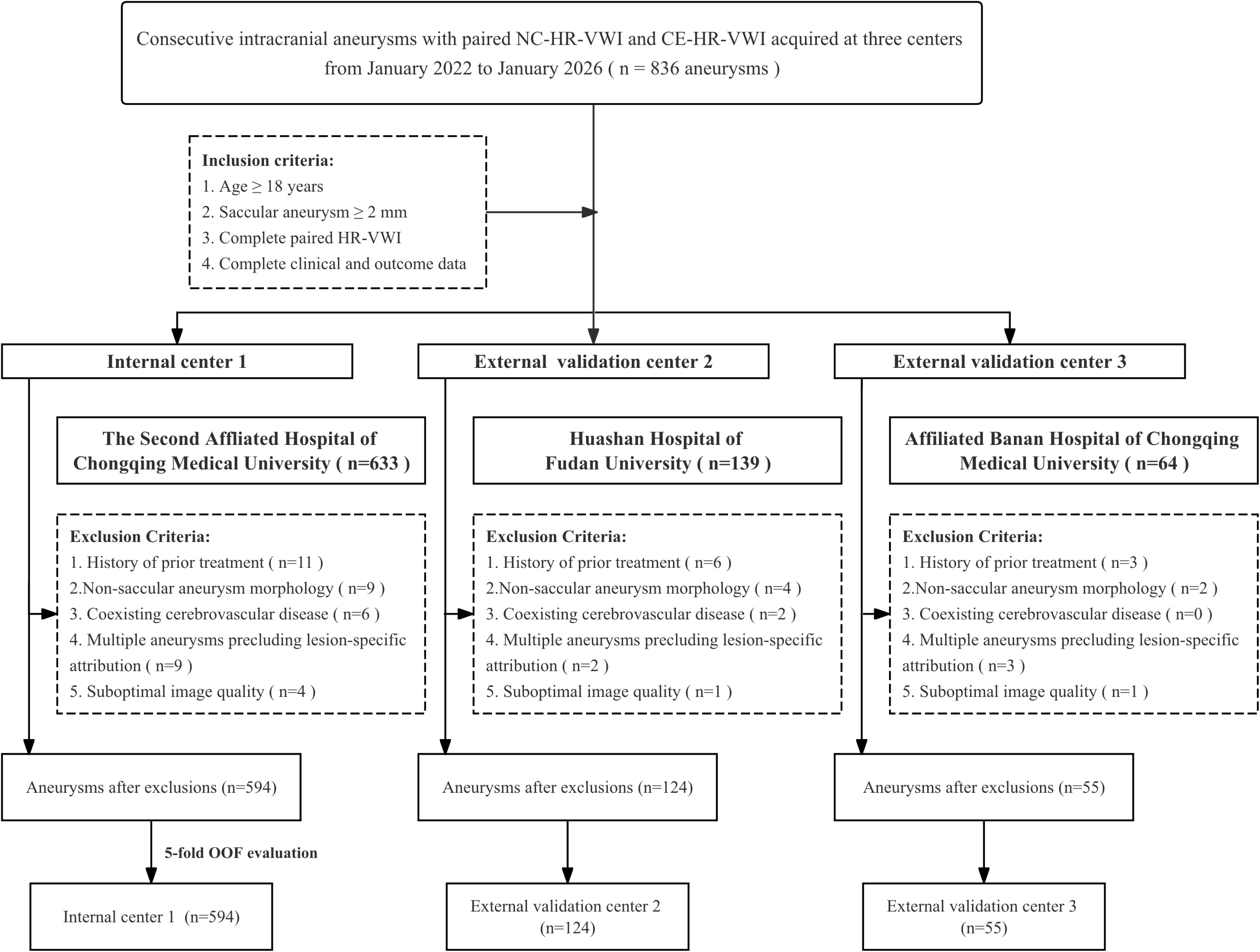
Patient enrollment flowchart. Screened records from three centers were assessed using common eligibility criteria; the final aneurysm cohorts underwent internal patient-level five-fold out-of-fold assessment or independent external validation. All n values represent aneurysms. HR-VWI, high-resolution vessel wall imaging; OOF, out-of-fold.

**Figure 2.**
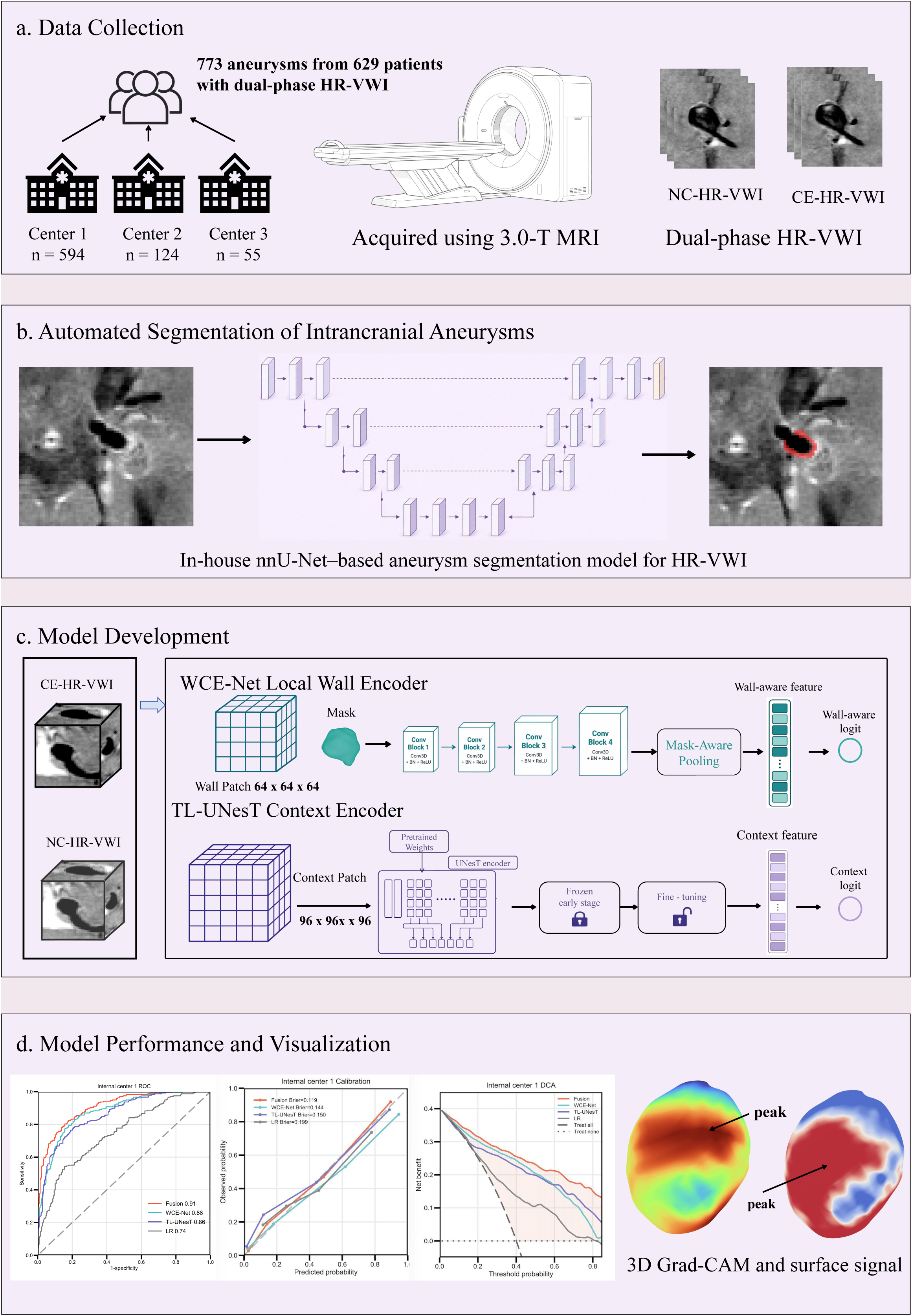
Overall study workflow. Panel a summarizes multicenter dual-phase HR-VWI acquisition; panel b, automated aneurysm segmentation; panel c, dual-phase local wall-context model development; and panel d, performance evaluation and surface visualization. CE, contrast-enhanced; DCA, decision curve analysis; HR-VWI, high-resolution vessel wall imaging; NC, non-contrast-enhanced; ROC, receiver operating characteristic. In panel c, WCE-Net and TL-UNesT were applied separately to NC and CE inputs, yielding four branch logits for final fusion.

After applying the inclusion and exclusion criteria, 629 patients with 773 intracranial aneurysms were included: 482 patients with 594 aneurysms from Center 1, 96 patients with 124 aneurysms from Center 2, and 51 patients with 55 aneurysms from Center 3. The unit of analysis was the aneurysm. For patients with multiple aneurysms, all aneurysms from the same patient were always assigned to the same dataset partition and the same cross-validation fold to avoid data leakage.

This multicenter retrospective study was reported in accordance with the 2024 update of the Checklist for Artificial Intelligence in Medical Imaging (CLAIM) and the STROBE statement for cohort studies.

### 2.2 Outcome Definition and Imaging Protocol

The primary endpoint was unstable intracranial aneurysm phenotype. Each aneurysm was classified as stable or unstable according to clinical status, follow-up changes, and imaging findings.

Stable aneurysms were defined as asymptomatic unruptured aneurysms without definite morphologic change during at least 1 year of follow-up. Unstable aneurysms were defined by any of the following criteria:

1. Ruptured aneurysms [17], presenting with subarachnoid hemorrhage;
2. Symptomatic unruptured aneurysms [17], defined as aneurysms associated with attributable clinical symptoms in the absence of evidence of subarachnoid hemorrhage, including cranial nerves II-VI palsy, sentinel headache, or focal neurologic deficit, with symptom onset within 2 weeks before presentation. Isolated nonspecific headache was not considered sufficient for classification as symptomatic unruptured aneurysm;
3. Growing unruptured aneurysms [17], defined as definite morphologic change on follow-up imaging, including de novo daughter sac formation, change in aneurysm shape, or an increase of more than 1 mm in maximum diameter.

For patients with multiple aneurysms, the responsible ruptured, symptomatic, or growing lesion was determined by integrating clinical records, hemorrhage distribution, aneurysm location, morphologic evolution, and imaging findings.

The unstable phenotype defined in this study was a composite endpoint based on clinical status, follow-up change, and imaging data, and model outputs were intended to identify aneurysms with this phenotype. The model task was binary classification of stable aneurysms versus aneurysms with a composite unstable phenotype; ruptured, symptomatic unruptured, and growing unruptured aneurysms were combined into one positive class and were not modeled as separate subtypes. All included cases underwent a standardized intracranial HR-VWI protocol. Detailed imaging parameters for each center are provided in Supplementary Table S1.

### 2.3 Aneurysm Segmentation and Image Preprocessing

Initial aneurysm masks were generated using an in-house automatic segmentation model based on nnU-Net and were reviewed and corrected when necessary in ITK-SNAP by two experienced neuroradiologists [18,19]; disagreements were adjudicated by a senior neuroradiologist. All subsequent preprocessing and model analyses were based on the final confirmed masks.

Image preprocessing included intensity normalization and mask binarization. For each HR-VWI image, intensities were clipped at the 0.5th and 99.5th percentiles and linearly normalized to the range of 0-1. Aneurysm masks were converted to binary masks. Two spatial scales of three-dimensional inputs were generated from the same aneurysm mask. For local wall inputs, the lesion region was localized using the three-dimensional mask bounding box, cropped after expansion by a fixed margin in each direction, and padded or cropped to 64 x 64 x 64; the corresponding image patch and binary mask were jointly input to the WCE-Net branch. For context inputs, a larger image patch centered on the mask centroid was cropped from the original image and resized to 96 x 96 x 96 for the transfer-learning UNesT branch. This workflow was applied separately to non-contrast and contrast-enhanced HR-VWI, resulting in four inputs for each aneurysm: non-contrast local wall, contrast-enhanced local wall, non-contrast context, and contrast-enhanced context.

### 2.4 Model Architecture and Development

We developed a WCE-Net-based dual-phase local wall-context fusion deep learning model. The model used non-contrast and contrast-enhanced HR-VWI as dual-phase inputs and constructed local wall branches and spatial context branches. Local wall branches extracted focal aneurysm wall imaging phenotypes, whereas context branches extracted broader perilesional spatial information. The final model consisted of four independent prediction branches: non-contrast WCE-Net local wall branch, contrast-enhanced WCE-Net local wall branch, non-contrast TL-UNesT context branch, and contrast-enhanced TL-UNesT context branch. The overall model architecture is shown in Figure 3.

**Figure 3.**
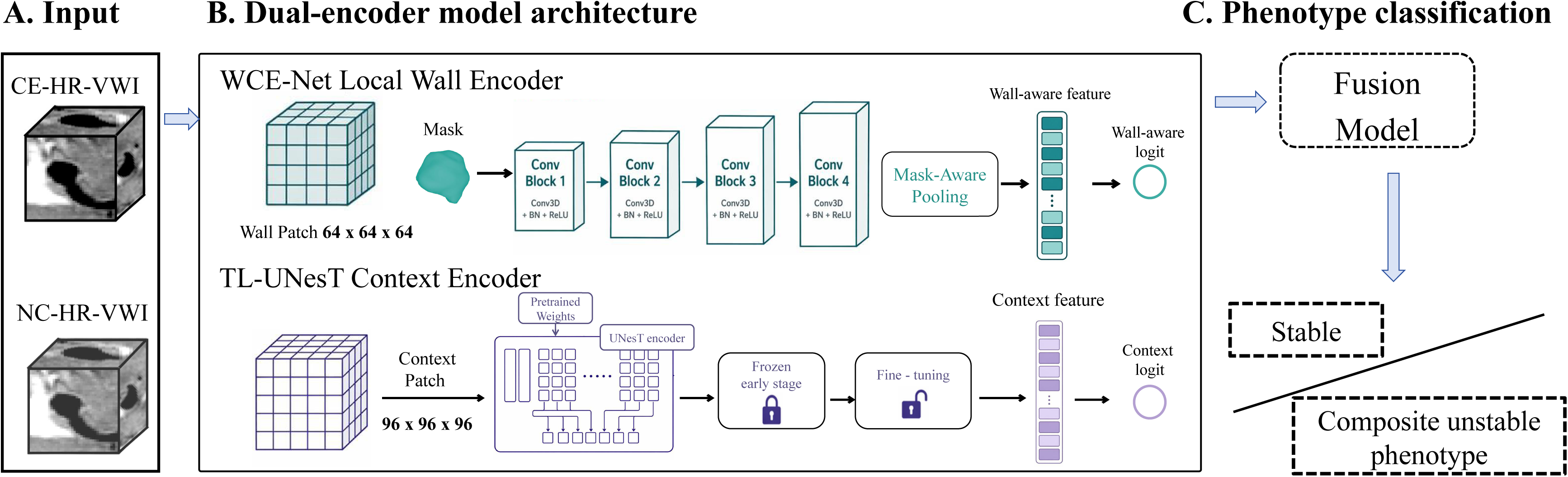
WCE-Net-based dual-encoder architecture. Panel a shows the dual-phase HR-VWI inputs; panel b shows the WCE-Net local wall and TL-UNesT context encoders; and panel c shows the fused risk output for stable versus composite unstable aneurysm phenotypes. HR-VWI, high-resolution vessel wall imaging; TL-UNesT, transfer-learning U-Net with Nested Transformers; WCE-Net, Wall-Constrained Encoding Network. Each encoder was applied separately to non-contrast and contrast-enhanced inputs, and the resulting four logits were combined by the fusion model.

WCE-Net was used for local wall feature extraction. Its inputs were 64 x 64 x 64 local wall image patches and corresponding binary masks, and the network architecture is summarized in Table 1. The network consisted of four three-dimensional convolutional modules and incorporated mask-constrained aggregation on the final feature map: the binary mask was interpolated to the same spatial size as the feature map and multiplied voxelwise by the feature map, and average pooling was performed only within the mask-covered region to obtain a local wall feature vector and output the local branch prediction logit.

**Table 1.** Architecture of WCE-Net.

| Module | Layer type | Output channels | Kernel size | Feature map change |
| --- | --- | --- | --- | --- |
| Input | Local image patch + mask | 1 | - | $64 \times 64 \times 64$ |
| Block 1 | Conv3D + BN + ReLU | 16 | $3 \times 3 \times 3 / 1$ | Spatial size preserved |
| Block 2 | Conv3D + BN + ReLU | 32 | $3 \times 3 \times 3 / 2$ | Downsampling |
| Block 3 | Conv3D + BN + ReLU | 64 | $3 \times 3 \times 3 / 2$ | Further downsampling |
| Block 4 | Conv3D + BN + ReLU | 128 | $3 \times 3 \times 3 / 1$ | Feature depth increased |
| Mask-aware pooling | Mask-guided weighted pooling | 128 | - | Collapsed to fixed-length vector |
| Classification head | Linear + ReLU + Dropout + Linear | 1 | - | Single logit output |

The context branch used transfer-learning-based 3D UNesT to extract spatial context features from 96 x 96 x 96 context patches. The UNesT backbone was initialized with the wholeBrainSeg Large UNEST segmentation pretrained weights from the MONAI Model Zoo and fine-tuned in the aneurysm context branch [15,20]. At the fusion stage, logit-level stacking was used: logits from the four branches were input to a logistic regression fuser to generate the final prediction. The logistic regression fuser, classification threshold, and temperature-scaling calibration parameters were estimated exclusively from aggregated five-fold OOF predictions in Center 1. No data from Centers 2 or 3 were used for model selection, fusion-weight estimation, threshold selection, or calibration [21].

### 2.5 Model Training and Local Encoder Comparison

All deep learning models were developed in Center 1 using five-fold cross-validation and internal OOF evaluation. Training used the AdamW optimizer [22] and weighted binary cross-entropy loss, with positive-class weights determined from the sample distribution of stable and unstable aneurysms in the internal training set. Automatic mixed precision and early stopping were used during training; the maximum number of epochs was 120, early-stopping patience was 15 epochs, and weight decay was 5 x 10^-4. All cross-validation splits were performed at the patient level; all aneurysms from the same patient were assigned to the same fold to prevent different aneurysms from one patient from entering both training and validation folds.

The initial learning rate of WCE-Net was 2 x 10^-4. For the transfer-learning context encoding branch, the 3D UNesT backbone loaded external pretrained weights [15,20]. During the first 10 epochs, backbone parameters were frozen and only the classification head was updated; the backbone was then unfrozen for joint fine-tuning. A grouped learning-rate strategy was used for the UNesT branch, with a learning rate of 2 x 10^-5 for the pretrained backbone and 2 x 10^-4 for the classification head.

To evaluate WCE-Net as a local wall encoder, we compared it with four commonly used three-dimensional encoders: ResNet34, DenseNet121, ConvNeXt, and MedNeXt [23–26]. All candidate local encoders used the same data splits, local wall patch inputs, label definition, and training strategy, and discrimination was evaluated in internal Center 1 and two independent external centers. To assess the effect of key model-design components on performance, we further performed wall-aware feature aggregation ablation and dual-phase input ablation; detailed results are provided in Supplementary Appendix 2 and Supplementary Tables S3-S4.

### 2.6 Model Performance Evaluation and Statistical Analysis

Discrimination was evaluated using receiver operating characteristic curves and the area under the curve (AUC). Accuracy, sensitivity, specificity, positive predictive value, and negative predictive value were calculated from confusion matrices at a fixed classification threshold. The threshold was determined as the optimal F1 threshold on aggregated five-fold OOF predictions from internal Center 1 and was fixed for external Centers 2 and 3. The 95% CIs for the primary model AUCs were estimated using 2000 patient-level cluster bootstrap resamples. Exploratory differences in AUC between models were estimated using patient-level clustered paired bootstrap resampling.

Calibration was evaluated using calibration curves, Brier score, and expected calibration error. Clinical utility was assessed with decision curve analysis [27]. Continuous variables are reported as mean +/-standard deviation or median (interquartile range), as appropriate, and categorical variables are reported as counts and percentages; between-group comparisons used appropriate parametric or nonparametric tests according to variable type and distribution. All statistical tests were two-sided, and P < 0.05 was considered statistically significant. Model development and statistical analyses were mainly performed in Python 3.9 using PyTorch [28], MONAI [29], scikit-learn [30], and related statistical packages. In addition, a clinical-imaging logistic regression model was constructed as a structured reference comparator based on clinical, morphologic, and aneurysm wall-related variables available across cohorts, and was compared with the deep learning models; details are provided in Supplementary Appendix 1, Supplementary Table S2, and Supplementary Figures S1–S3.

### 2.7 Anatomical Interpretability Analysis

To assess the spatial distribution of model-prediction-related regions on the aneurysm wall, we performed three-dimensional Grad-CAM analysis on the WCE-Net local wall branches [16]. Because WCE-Net directly receives local wall image patches and corresponding binary masks, its attention responses can be aligned with aneurysm wall locations. Grad-CAM was generated separately for non-contrast and contrast-enhanced local wall inputs. Using the branch prediction logit as the target, gradient weights of the final convolutional feature map were calculated to obtain three-dimensional attention response maps.

To obtain visualizations more consistent with aneurysm wall anatomy, three-dimensional Grad-CAM responses were further projected onto the aneurysm wall surface. Specifically, the local image patch, binary mask, and Grad-CAM response map were maintained in the same spatial coordinate system. After mild smoothing of the aneurysm mask, a three-dimensional wall mesh was reconstructed using the marching cubes algorithm [31]; small fragments disconnected from the main component were removed, leaving only the main aneurysm surface. Grad-CAM response values and HR-VWI signal intensities were then sampled at each vertex of the same surface mesh to generate a surface attention map and a surface signal map. Because both types of information were displayed on the same wall-surface geometry, they could be used to visually compare the spatial relationship between model-attended regions and high wall-signal regions and to demonstrate the continuity and focal distribution of model attention on the aneurysm wall from a three-dimensional perspective.

To quantitatively describe the correspondence between model attention and wall-signal distribution, we further performed surface hotspot overlap analysis. The top 10% of surface vertices with the highest Grad-CAM responses were defined as model-attention hotspots, and the top 10% of vertices with the highest HR-VWI signal intensity were defined as high wall-signal hotspots. Dice coefficients between the two hotspot regions were calculated. Pearson and Spearman correlation coefficients between surface Grad-CAM responses and surface signal intensities were also calculated to assess spatial consistency. For the quantitative surface analysis, Grad-CAM maps from the non-contrast WCE-Net branch were evaluated in all 773 included aneurysms.

## 3. Results

### 3.1 Patient Characteristics

A total of 629 patients with 773 intracranial aneurysms were included, comprising 594, 124, and 55 aneurysms in Centers 1, 2, and 3, respectively. The proportions of unstable aneurysms were similar across the three centers, at 40.1%, 40.3%, and 40.0%, respectively. Clinical and imaging characteristics of the cohorts are shown in Table 2.

**Table 2.** Baseline characteristics of intracranial aneurysms across three cohorts.

|  | Internal development cohort (n = 594) |  |  | External validation Center 2 (n = 124) |  |  | External validation Center 3 (n = 55) |  |  |
| --- | --- | --- | --- | --- | --- | --- | --- | --- | --- |
| Characteristic | Stable (n = 356) | Unstable (n = 238) | *P* | Stable (n = 74) | Unstable (n = 50) | *P* | Stable (n = 33) | Unstable (n = 22) | *P* |
| Age (years) | 61.0 (54.0–69.0) | 59.5 (51.0–67.0) | 0.017 | 57.0 (51.0–63.5) | 59.0 (53.0–66.0) | 0.338 | 56.0 (48.0–60.0) | 56.0 (46.2–60.0) | 0.546 |
| Male sex | 111 (31.2) | 87 (36.6) | 0.203 | 22 (29.7) | 16 (32.0) | 0.944 | 9 (27.3) | 10 (45.5) | 0.271 |
| Hypertension | 185 (52.0) | 136 (57.1) | 0.247 | 49 (66.2) | 27 (54.0) | 0.237 | 22 (66.7) | 12 (54.5) | 0.533 |
| Diabetes mellitus | 59 (16.6) | 25 (10.5) | 0.050 | 14 (18.9) | 10 (20.0) | 1.000 | 7 (21.2) | 5 (22.7) | 1.000 |
| Coronary artery disease | 45 (12.6) | 16 (6.7) | 0.028 | 11 (14.9) | 21 (42.0) | 0.001 | 3 (9.1) | 10 (45.5) | 0.005 |
| Smoking history | 84 (23.6) | 61 (25.6) | 0.640 | 13 (17.6) | 10 (20.0) | 0.915 | 7 (21.2) | 3 (13.6) | 0.723 |
| Maximum diameter (mm) | 4.02 (3.10–5.15) | 5.25 (3.60–7.84) | < 0.001 | 5.44 (3.96–6.82) | 9.13 (5.59–17.04) | < 0.001 | 4.54 (3.95–7.14) | 7.35 (5.58–10.23) | 0.005 |
| Aneurysm size group |  |  | < 0.001 |  |  | < 0.001 |  |  | 0.005 |
| < 5 mm | 259 (72.8) | 113 (47.5) |  | 36 (48.6) | 10 (20.0) |  | 19 (57.6) | 3 (13.6) |  |
| 5–10 mm | 88 (24.7) | 81 (34.0) |  | 29 (39.2) | 20 (40.0) |  | 8 (24.2) | 12 (54.5) |  |
| > 10 mm | 9 (2.5) | 44 (18.5) |  | 9 (12.2) | 20 (40.0) |  | 6 (18.2) | 7 (31.8) |  |
| Location |  |  | < 0.001 |  |  | 0.262 |  |  | 0.153 |
| ICA | 239 (67.1) | 111 (46.6) |  | 39 (52.7) | 23 (46.0) |  | 17 (51.5) | 8 (36.4) |  |
| MCA | 51 (14.3) | 51 (21.4) |  | 15 (20.3) | 17 (34.0) |  | 11 (33.3) | 7 (31.8) |  |
| ACA/ACoM | 35 (9.8) | 34 (14.3) |  | 13 (17.6) | 6 (12.0) |  | 5 (15.2) | 3 (13.6) |  |
| PComA | 18 (5.1) | 24 (10.1) |  | 5 (6.8) | 1 (2.0) |  | 0 (0.0) | 1 (4.5) |  |
| Posterior circulation | 13 (3.7) | 18 (7.6) |  | 2 (2.7) | 3 (6.0) |  | 0 (0.0) | 3 (13.6) |  |

Data are presented as median (interquartile range) for continuous variables or n (%) for categorical variables. The unit of analysis is the aneurysm. Continuous variables were compared using the Mann–Whitney U test. Categorical variables were compared using the chi-square test or the Fisher exact test when any expected cell count was less than 5.
ICA, internal carotid artery; MCA, middle cerebral artery; ACA, anterior cerebral artery; ACoM, anterior communicating artery; PComA, posterior communicating artery.

### 3.2 Discriminative Performance of Local Encoders and the Final Fusion Model

ROC performance of the five three-dimensional local encoders is shown in Figure 4a-c. WCE-Net achieved the highest AUCs in internal Center 1, external Center 2, and external Center 3, with values of 0.878, 0.848, and 0.844, respectively; the AUC ranges of the other encoders in the three cohorts were 0.674-0.790, 0.634-0.759, and 0.656-0.771, respectively. ROC performance of the final dual-phase local wall-context fusion model, WCE-Net alone, TL-UNesT alone, and the clinical-imaging logistic regression baseline is shown in Figure 4d-f.

**Figure 4.**
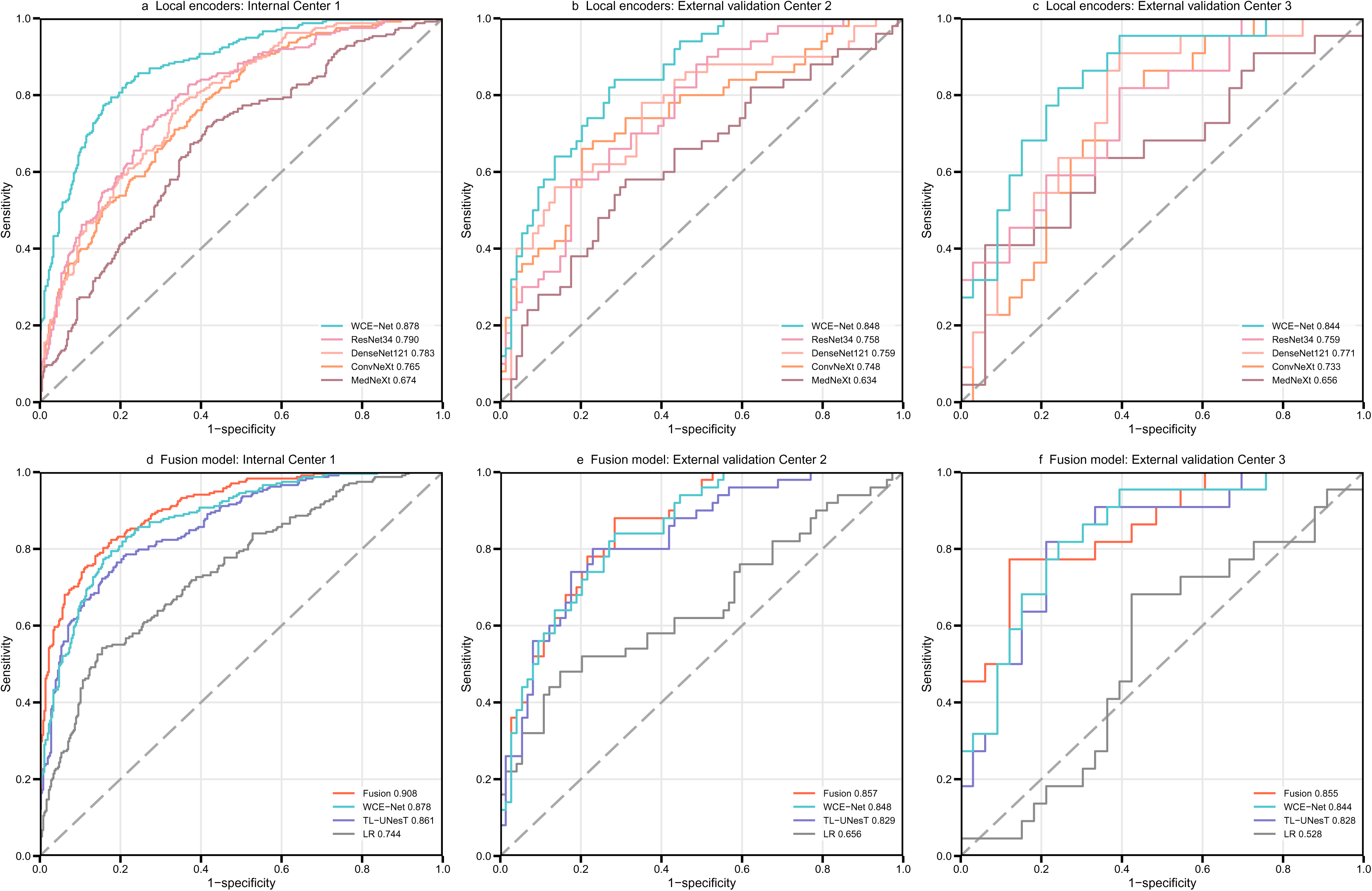
Receiver operating characteristic curves for candidate local encoders and the final fusion model. Panels a-c show ROC curves for WCE-Net, ResNet34, DenseNet121, ConvNeXt, and MedNeXt in internal Center 1, external validation Center 2, and external validation Center 3. Panels d-f show ROC curves for the final fusion model, WCE-Net, TL-UNesT, and logistic regression in the same cohorts. AUC values are shown in the legends. LR = logistic regression.

The final fusion model showed favorable discriminative performance in the internal assessment and both external validation cohorts (Figure 4, Table 3). AUCs in Center 1, external Center 2, and external Center 3 were 0.908, 0.857, and 0.855, respectively, and were numerically higher than those of the WCE-Net branch alone, the TL-UNesT branch alone, and the logistic regression baseline. The clinical-imaging logistic regression baseline was used as a structured reference comparator based on variables available across cohorts, and its cross-center performance should be interpreted in this context. Complete classification metrics are shown in Table 3.

**Table 3.**
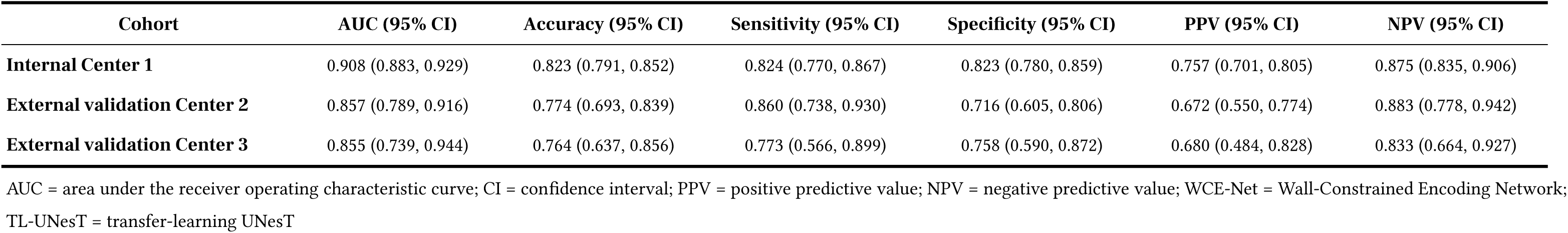
Predictive performance of the final fusion model in the internal center and external validation centers.

| Cohort | AUC (95% CI) | Accuracy (95% CI) | Sensitivity (95% CI) | Specificity (95% CI) | PPV (95% CI) | NPV (95% CI) |
| --- | --- | --- | --- | --- | --- | --- |
| Internal Center 1 | 0.908 (0.883, 0.929) | 0.823 (0.791, 0.852) | 0.824 (0.770, 0.867) | 0.823 (0.780, 0.859) | 0.757 (0.701, 0.805) | 0.875 (0.835, 0.906) |
| External validation Center 2 | 0.857 (0.789, 0.916) | 0.774 (0.693, 0.839) | 0.860 (0.738, 0.930) | 0.716 (0.605, 0.806) | 0.672 (0.550, 0.774) | 0.883 (0.778, 0.942) |
| External validation Center 3 | 0.855 (0.739, 0.944) | 0.764 (0.637, 0.856) | 0.773 (0.566, 0.899) | 0.758 (0.590, 0.872) | 0.680 (0.484, 0.828) | 0.833 (0.664, 0.927) |

AUC = area under the receiver operating characteristic curve; CI = confidence interval; PPV = positive predictive value; NPV = negative predictive value; WCE-Net = Wall-Constrained Encoding Network; TL-UNesT = transfer-learning UNesT

### 3.3 Calibration and Clinical Net Benefit

Calibration curves and decision curve analysis results are shown in Figure 5. In calibration analysis, Brier scores of the fusion model in internal Center 1, external Center 2, and external Center 3 were 0.119, 0.153, and 0.150, respectively. Compared with the WCE-Net branch alone and the TL-UNesT branch alone, the fusion model achieved the lowest Brier score in all three validation cohorts. In decision curve analysis, the fusion model showed higher net benefit across most threshold-probability ranges in the internal and external centers and overall outperformed WCE-Net and TL-UNesT alone.

**Figure 5.**
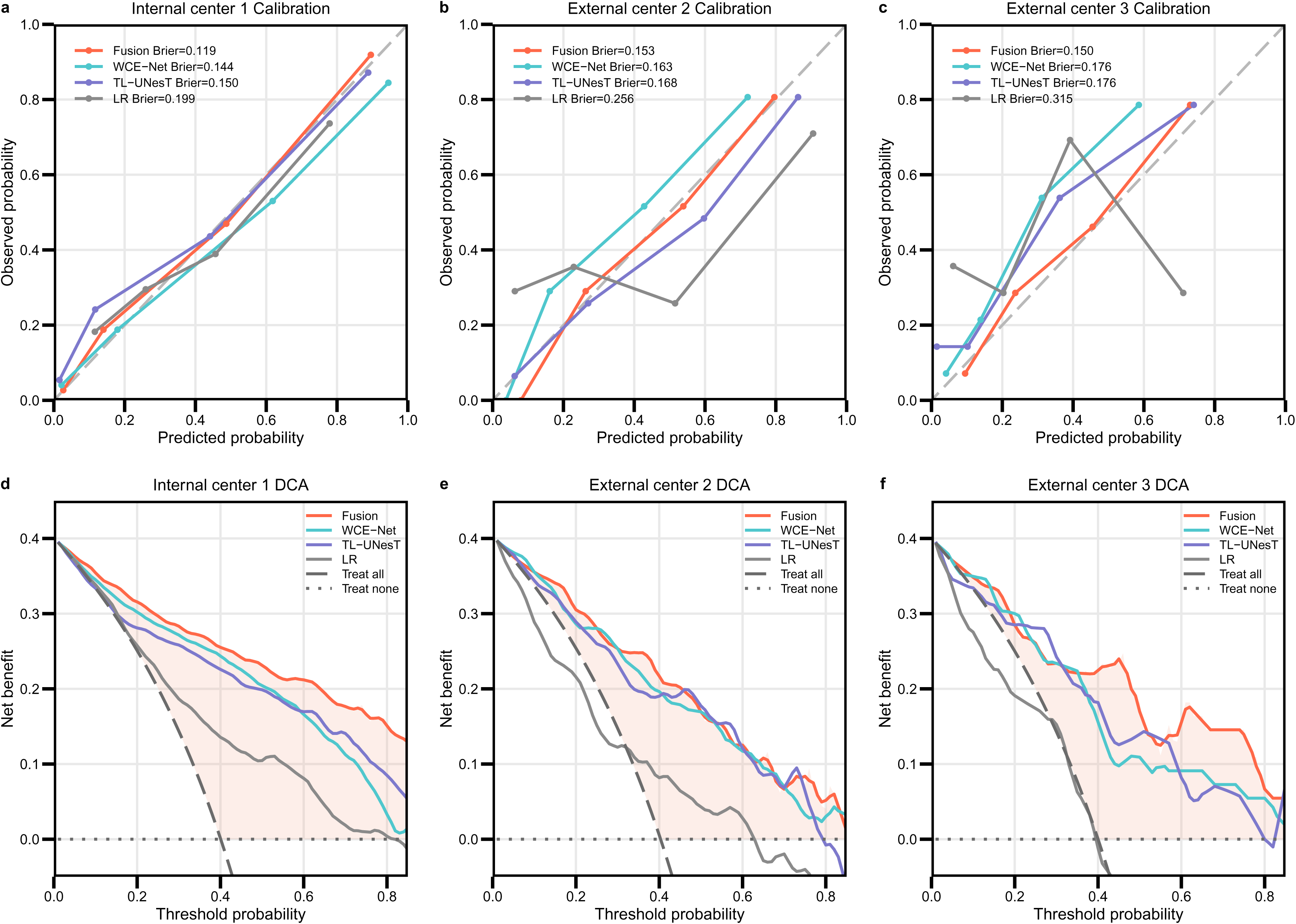
Calibration and decision curves for deep learning and clinical-imaging models. Panels a-c show calibration curves and panels d-f show decision curve analyses for the internal center and two external validation centers, with the clinical-imaging logistic regression model as a reference comparator. The fusion model achieved Brier scores of 0.119, 0.153, and 0.150, respectively. WCE-Net achieved Brier scores of 0.144, 0.163, and 0.176, whereas TL-UNesT achieved Brier scores of 0.150, 0.168, and 0.176, respectively. Because external validation Center 3 included a relatively small number of aneurysms, its calibration and decision curve patterns should be interpreted in the context of sample size.

### 3.4 Three-Dimensional Grad-CAM Surface Visualization

Two-dimensional and three-dimensional Grad-CAM visualization results are shown in Figure 6 and Supplementary Figure S4. Two-dimensional Grad-CAM heatmaps showed that, in representative unstable aneurysms, model attention was mainly concentrated on the aneurysm wall and adjacent regions rather than being diffusely distributed in surrounding background structures. For stable aneurysms, Grad-CAM responses were generally weaker or more diffuse. These two-dimensional heatmaps provided an intuitive slice-level visualization of model-attended regions.

**Figure 6.**
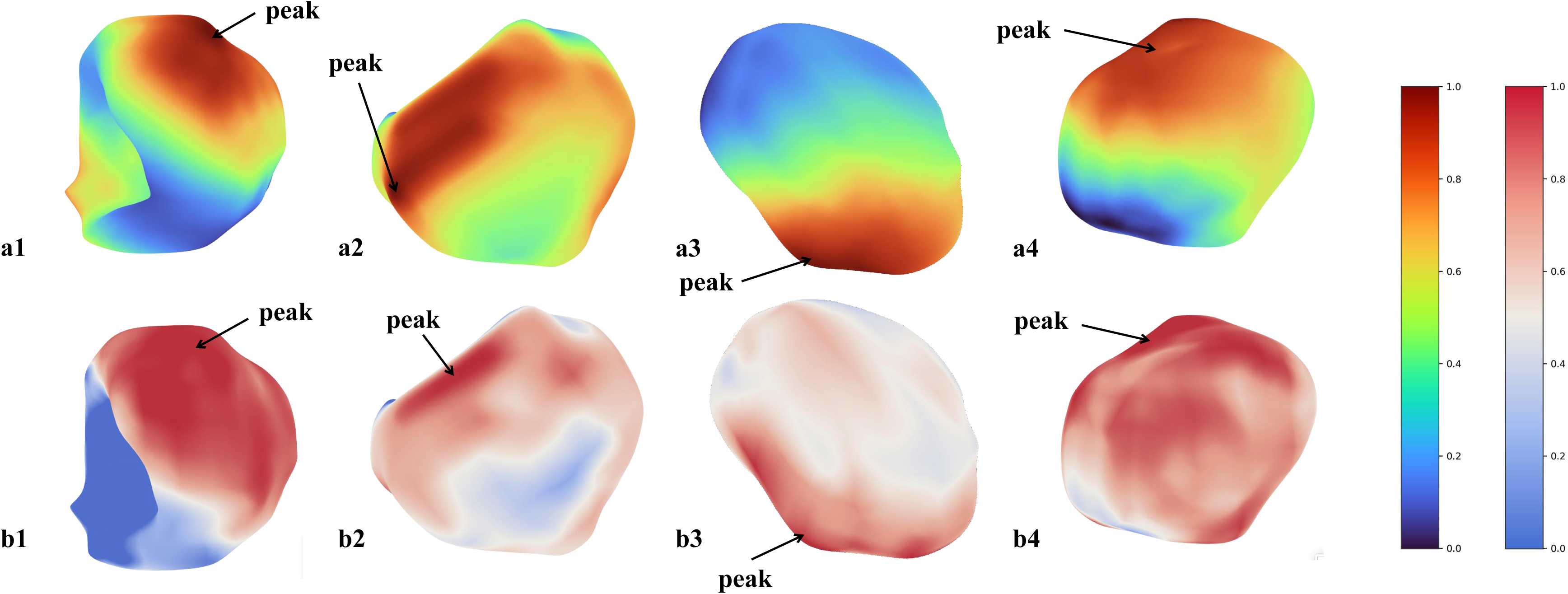
Three-dimensional surface Grad-CAM visualization. Panels a1-a4 show surface Grad-CAM maps, and panels b1-b4 show the corresponding HR-VWI surface-signal maps. Within each column, both maps are displayed on the same aneurysm wall geometry; arrows mark peak values and warmer colors indicate higher normalized values.

In the overall surface hotspot overlap analysis, the two types of hotspots showed partial spatial overlap. Across all cases, the mean and median Top 10% Dice coefficients were 0.430 and 0.437, respectively. Pointwise correlation analysis further showed a moderate positive correlation between surface Grad-CAM responses and surface signal intensities; mean and median Pearson correlation coefficients were 0.541 and 0.591, respectively, and mean and median Spearman correlation coefficients were 0.504 and 0.543, respectively.

## 4. Discussion

In this multicenter retrospective study, we developed and evaluated a WCE-Net-based wall-focused deep learning framework for identifying composite unstable intracranial aneurysm phenotypes on dual-phase HR-VWI. By using mask-constrained feature aggregation, WCE-Net was designed to extract local wall-related imaging information from small, thin, and irregular aneurysm wall regions. After integrating local wall features, spatial context information, and dual-phase HR-VWI phenotypes, the final fusion model showed favorable discrimination in internal out-of-fold assessment and two independent external validation cohorts. In addition, three-dimensional surface Grad-CAM mapped model attention onto the reconstructed aneurysm wall surface, providing an anatomically referenced visualization of model output.

HR-VWI has been recognized as an important imaging tool for characterizing unstable intracranial aneurysms, but aneurysm walls are typically small, thin, and irregular in three-dimensional images [4]. In this setting, nonlesional background signal can account for a large proportion of local image patches, and direct use of general-purpose three-dimensional encoders may be insufficient for stable aggregation of effective features originating from the aneurysm wall [23–26]. WCE-Net was designed for this task-specific scenario: it uses a lightweight three-dimensional convolutional structure and mask-guided feature aggregation to concentrate feature representation on the aneurysm wall region. In this study, WCE-Net achieved higher AUCs than the compared general-purpose three-dimensional encoders in both internal and external validation cohorts. This suggests that an anatomy-oriented, task-specific design may improve the efficiency with which models use local wall phenotypes in HR-VWI-based aneurysm wall modeling. For small-volume medical imaging tasks with a clear anatomical target, model design may depend not only on increasing network complexity but also on aligning the encoder architecture with modality-specific features and the target anatomical region.

This study introduced a transfer-learning UNesT context encoder to extract spatial context information from larger three-dimensional image patches and fuse it with WCE-Net local wall features. The fusion model achieved higher AUCs than WCE-Net or TL-UNesT alone in all three cohorts, supporting complementary value among local wall information, spatial context information, and dual-phase HR-VWI phenotypes. Information captured by the spatial context branch may include not only perianeurysmal anatomical relationships but also indirect imaging factors associated with unstable phenotypes, such as aneurysm size, morphologic complexity, and adjacent vascular structures [5,6,8]. Therefore, the improved performance of the fusion model should not be interpreted simply as the effect of a single wall-enhancement signal. Rather, the framework likely integrates two complementary categories of information: local aneurysm wall phenotypes and perilesional spatial phenotypes.

The main value of three-dimensional surface Grad-CAM is that it visualizes model attention using the aneurysm wall as the anatomical reference. Compared with conventional two-dimensional slice heatmaps, surface projection displays model-attention distribution and HR-VWI surface signal distribution on the same wall-surface geometry [16], enabling more intuitive assessment of their spatial correspondence. In this study, Grad-CAM hotspots showed some consistency with wall imaging phenotypes. However, Grad-CAM remains a post hoc visualization method; it cannot prove that model-attended regions correspond to true pathological changes and cannot replace histological validation [32,33].

This study has several limitations. First, although two independent external validation cohorts were included, the retrospective design requires prospective validation to assess generalizability, clinical utility, and impact on treatment decisions. Second, Center 3 had a relatively small sample size; therefore, its performance estimates, calibration, and decision curve results should be interpreted cautiously. Third, the endpoint was a composite unstable phenotype including ruptured, symptomatic unruptured, and growing aneurysms, which may represent biologically heterogeneous subtypes. Thus, the model should not be interpreted as directly predicting future rupture risk in unruptured aneurysms. Fourth, surface Grad-CAM is a post hoc visualization method and cannot prove pathological causality or replace histological validation. Future studies should evaluate this framework in larger prospective cohorts and explore subtype-specific and longitudinal prediction tasks.

In conclusion, this study proposed a WCE-Net-based wall-focused deep learning framework for identifying composite unstable intracranial aneurysm phenotypes on dual-phase HR-VWI. The framework integrates local aneurysm wall features, spatial context information, and dual-phase imaging phenotypes and showed favorable discrimination in a multicenter retrospective cohort. Three-dimensional surface Grad-CAM provided anatomical visualization of model attention referenced to the aneurysm wall. This framework may provide a technical approach for HR-VWI-based imaging phenotype assessment of intracranial aneurysms.

## Data Availability

The code and datasets generated and/or analysed during the current study are not publicly available due to patient privacy and institutional restrictions but may be available from the corresponding author on reasonable request and subject to institutional approval.

## Declarations

### Ethics approval and consent to participate

This multicentre retrospective study was approved by the Ethics Committee of the Second Affiliated Hospital of Chongqing Medical University (approval/reference No. 194) and was conducted in accordance with the Declaration of Helsinki and its subsequent amendments. The requirement for written informed consent was waived by the ethics committee due to the retrospective nature of the study and the use of anonymised/de-identified clinical and imaging data.

### Consent for publication

Not applicable.

### Authors’ contributions

WY and ZW contributed equally to this work and share first authorship. WY and ZW contributed to study design, data collection, model development, data analysis, and drafting of the manuscript. QW contributed to data collection, image processing, and model evaluation. XH contributed to data curation, statistical analysis, and result interpretation. JT and XW contributed to data collection, imaging data organisation, and manuscript revision. RL contributed to image preprocessing, data management, and technical validation. YY contributed to clinical data interpretation and manuscript revision. DW and GW contributed to external data collection/validation and manuscript revision. TC conceived and supervised the study, contributed to study design, interpreted the results, critically revised the manuscript, and is the corresponding author. All authors read and approved the final manuscript.

## Abbreviations

ACA: anterior cerebral artery
AComA: anterior communicating artery
AdamW: Adam with decoupled weight decay
AUC: area under the receiver operating characteristic curve
BN: batch normalization
CE: contrast-enhanced
CI: confidence interval
Conv3D: three-dimensional convolution
DCA: decision curve analysis
ECE: expected calibration error
Grad-CAM: gradient-weighted class activation mapping
HR-VWI: high-resolution vessel wall imaging
ICA: internal carotid artery
LR: logistic regression
MCA: middle cerebral artery
MRI: magnetic resonance imaging
NC: non-contrast-enhanced
NPV: negative predictive value
OOF: out-of-fold
PComA: posterior communicating artery
PPV: positive predictive value
ReLU: rectified linear unit
ROC: receiver operating characteristic
TL-UNesT: transfer-learning UNesT
TOF-MRA: time-of-flight magnetic resonance angiography
UNesT: U-Net with Nested Transformers
WCE-Net: Wall-Constrained Encoding Network

## Acknowledgements

Not applicable.

## Authors’ information (optional)

Not applicable.

## Supplementary Material

**Supplementary Table S1.**
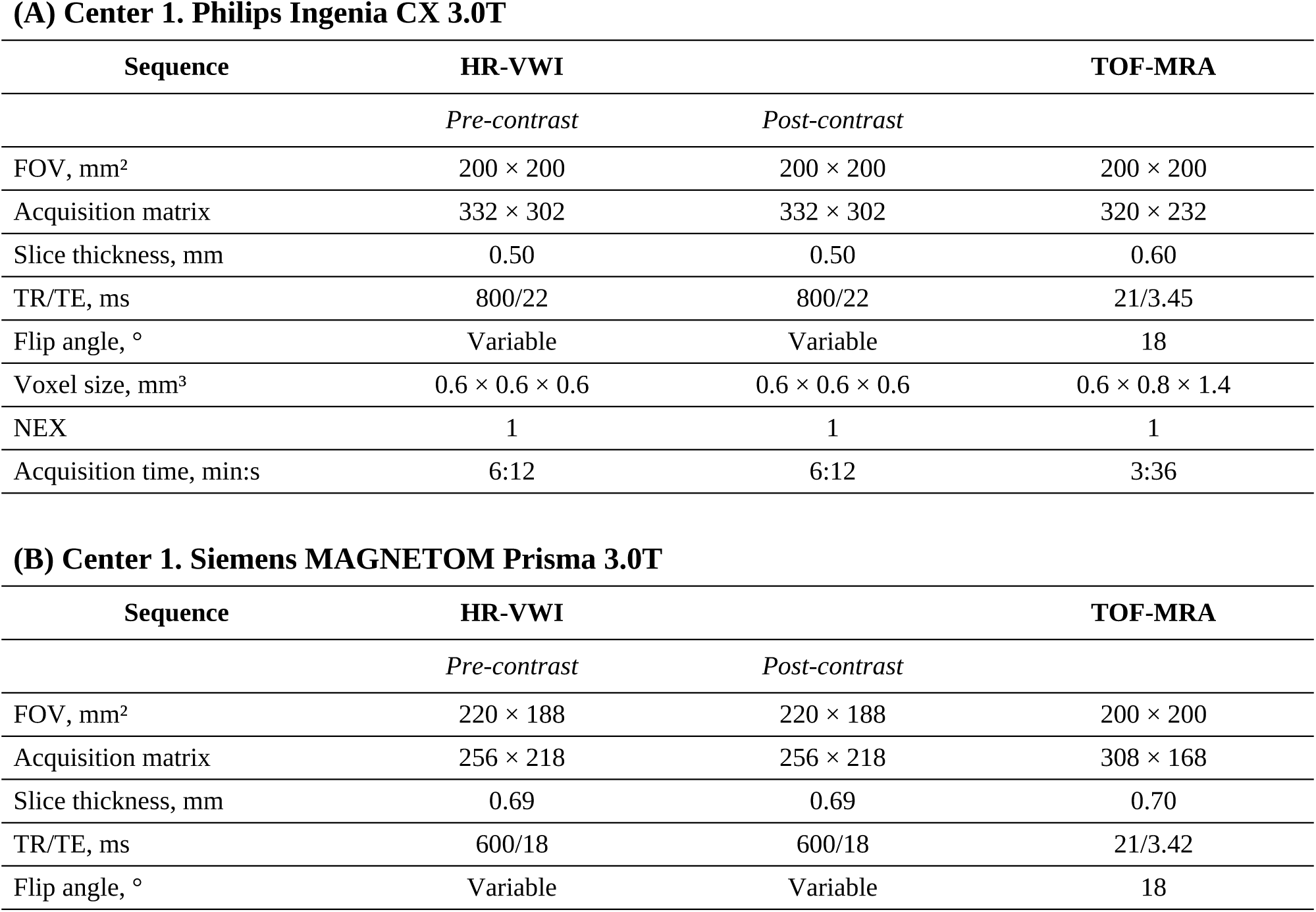
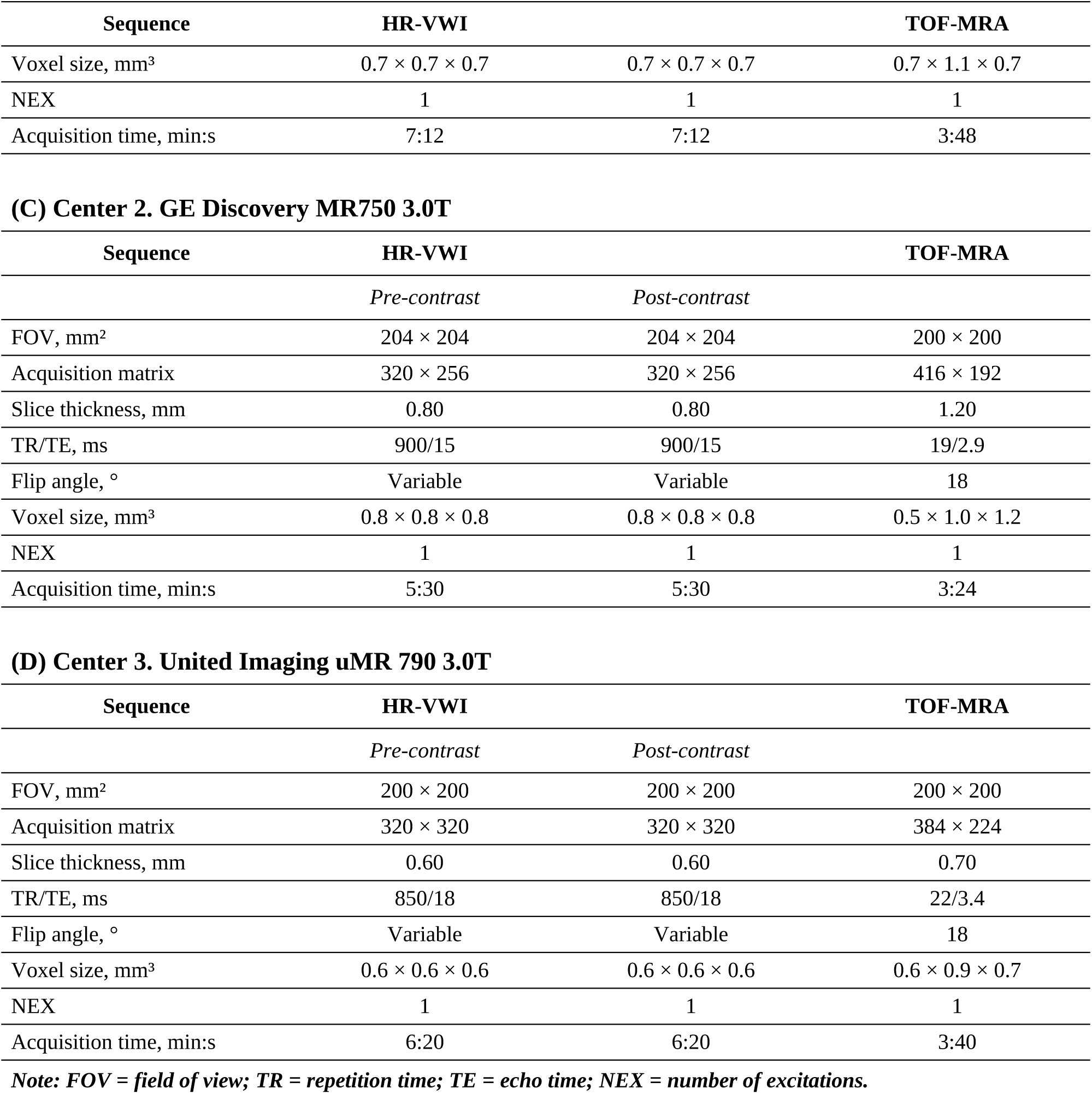
Scan parameters of dual-phase HR-VWI and TOF-MRA across participating cohorts. Imaging was performed at three centers using the following 3.0-T MRI scanners: Center 1, Ingenia CX (Philips Healthcare, Best, the Netherlands) with a 32-channel head coil and MAGNETOM Prisma (Siemens Healthineers, Erlangen, Germany) with a 64-channel head-neck coil; Center 2, Discovery MR750 (GE Healthcare, Milwaukee, WI, USA) with a 32-channel head coil; and Center 3, United Imaging uMR 790 (United Imaging Healthcare, Shanghai, China). All centers acquired three-dimensional TOF-MRA followed by three-dimensional black-blood T1-weighted HR-VWI. HR-VWI was performed using vendor-specific three-dimensional black-blood T1-weighted sequences in the corresponding cohorts. Post-contrast HR-VWI (CE-HR-VWI) was acquired approximately 5 min after intravenous administration of gadoterate meglumine (Gd-DOTA; 0.1 mmol/kg; Jiangsu Hengrui Pharmaceuticals Co., Ltd.) using the same sequence parameters as pre-contrast HR-VWI. Although acquisition parameters varied across cohorts, all scans followed standardized dual-phase HR-VWI protocols routinely used for intracranial aneurysm assessment. Detailed scan parameters for each center are listed below.

## Supplementary Appendix 1. Clinical-imaging reference model

To provide a structured clinical-imaging reference model for comparison with the deep learning models and to explore the potential incremental value of deep learning beyond available structured variables, we developed a logistic regression reference model. The internal cohort included 594 aneurysms from Center 1, and the external validation cohorts included 124 aneurysms from Center 2 and 55 aneurysms from Center 3. The reference outcome was the binary composite instability endpoint used in the main study. Internal performance was evaluated using patient-level five-fold out-of-fold validation.

Candidate variables included structured clinical, morphologic, scoring, and aneurysm wall-related variables available across all cohorts. These variables included age, sex, hypertension, hyperlipidemia, coronary artery disease, diabetes mellitus, smoking history, aneurysm location, irregular shape, daughter sac, neck width, transverse diameter, height, maximum diameter, sidewall or bifurcation type, size ratio, dome-to-neck ratio, aneurysm wall thickness, wall uniformity, thrombus or intramural hematoma, wall enhancement index, CRstalk, enhancement ratio, intracranial and parent artery atherosclerosis, and neck-to-parent ratio.

The location variable was one-hot encoded, and the remaining variables were treated as numeric variables. Imputation and standardization parameters were estimated within the corresponding training fold and then applied to the validation fold to avoid information leakage. The logistic regression model was fitted with L2 regularization. Model performance in the internal cohort was calculated from aggregated five-fold out-of-fold predictions. For external validation, the model was retrained on the full internal cohort and then directly applied to the two external validation cohorts.

The classification threshold was determined according to the maximum Youden index from the internal out-of-fold predicted probabilities and was fixed for internal performance summary and external validation. Model performance was evaluated using AUC, accuracy, sensitivity, specificity, positive predictive value, negative predictive value, Brier score, calibration curves, and decision curve analysis. This baseline model was further compared with WCE-Net, TL-UNesT, and the final fusion model.

**Supplementary Table S2.** Performance of the clinical-imaging logistic regression reference model.

| Cohort | AUC (95% CI) | Accuracy (95% CI) | Sensitivity (95% CI) | Specificity (95% CI) | PPV (95% CI) | NPV (95% CI) | Brier score |
| --- | --- | --- | --- | --- | --- | --- | --- |
| Internal Center 1 | 0.744 (0.704, 0.783) | 0.724 (0.689, 0.759) | 0.542 (0.480, 0.608) | 0.846 (0.805, 0.883) | 0.701 (0.634, 0.768) | 0.734 (0.691, 0.776) | 0.199 |
| External validation Center 2 | 0.656 (0.552, 0.756) | 0.645 (0.556, 0.726) | 0.520 (0.383, 0.659) | 0.730 (0.627, 0.831) | 0.565 (0.421, 0.708) | 0.692 (0.585, 0.792) | 0.256 |
| External validation Center 3 | 0.528 (0.369, 0.682) | 0.545 (0.400, 0.673) | 0.182 (0.042, 0.364) | 0.788 (0.643, 0.923) | 0.364 (0.100, 0.667) | 0.591 (0.435, 0.732) | 0.315 |

Abbreviations: CI = confidence interval; PPV = positive predictive value; NPV = negative predictive value.

**Supplementary Figure S1.**
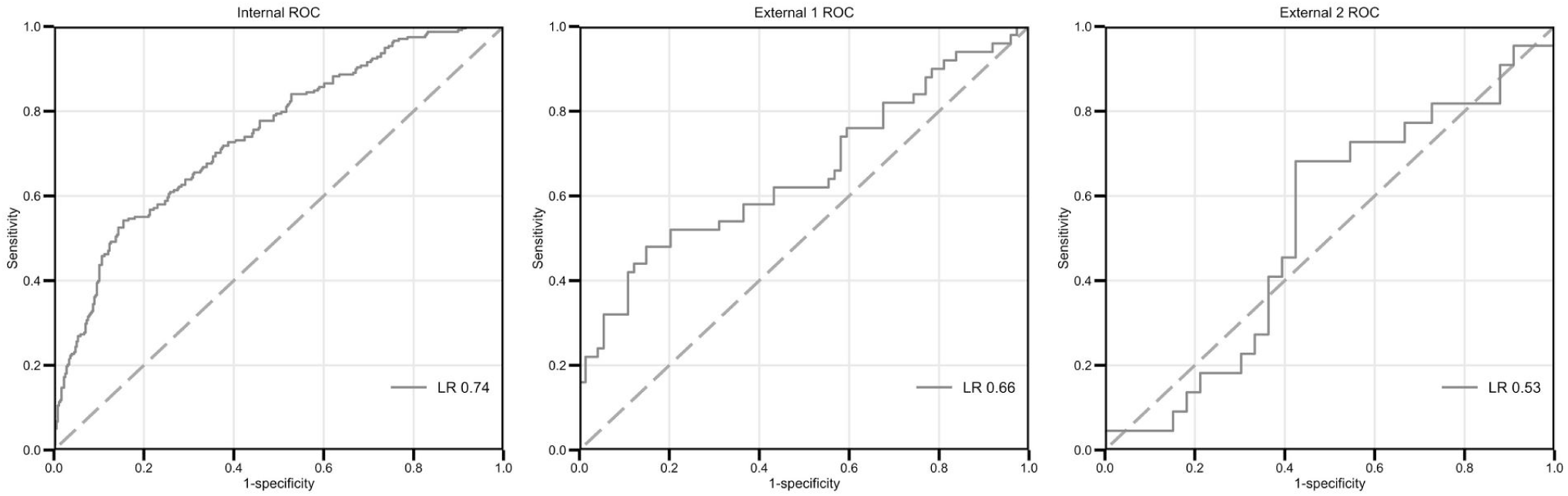
ROC curves of the clinical-imaging logistic regression reference model in the internal cohort and two external validation cohorts.

**Supplementary Figure S2.**
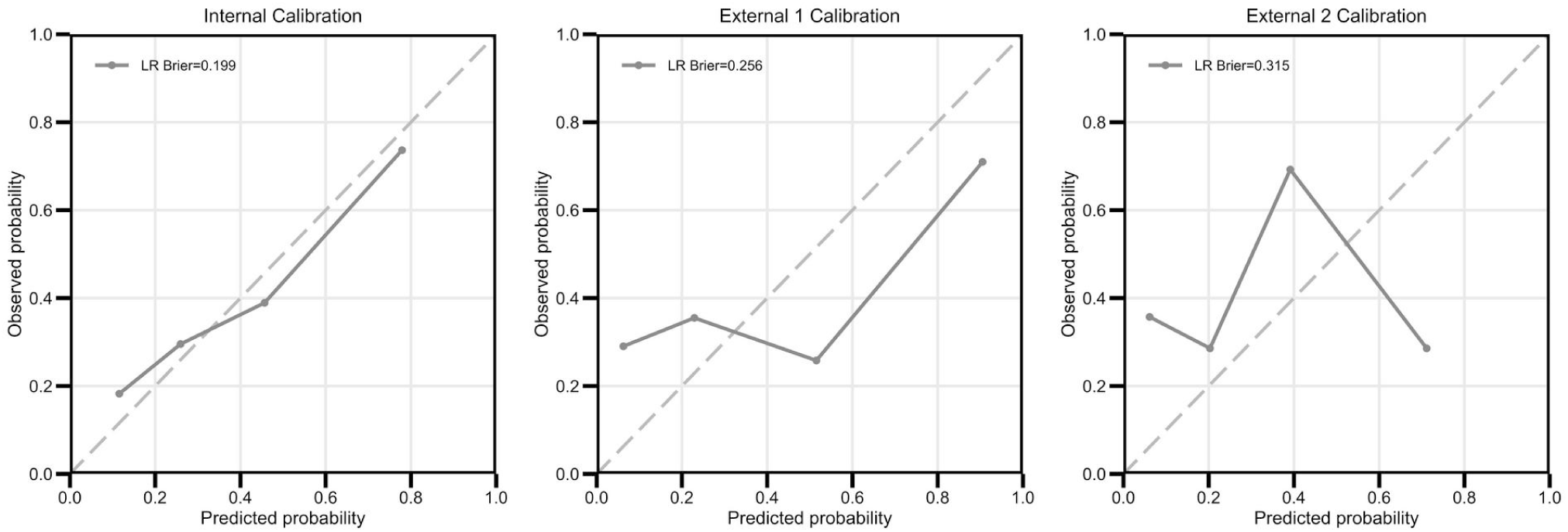
Calibration curves of the clinical-imaging logistic regression reference model in the internal cohort and two external validation cohorts.

**Supplementary Figure S3.**
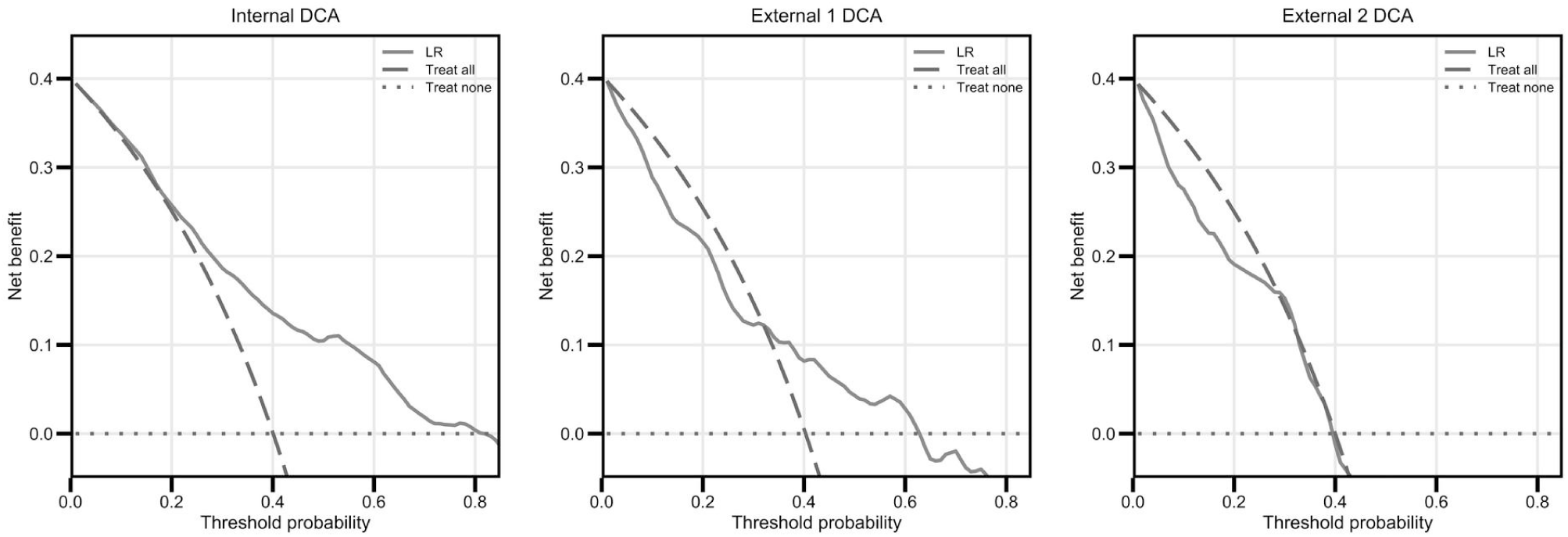
Decision curve analysis of the clinical-imaging logistic regression reference model in the internal cohort and two external validation cohorts.

**Supplementary Figure S4.**
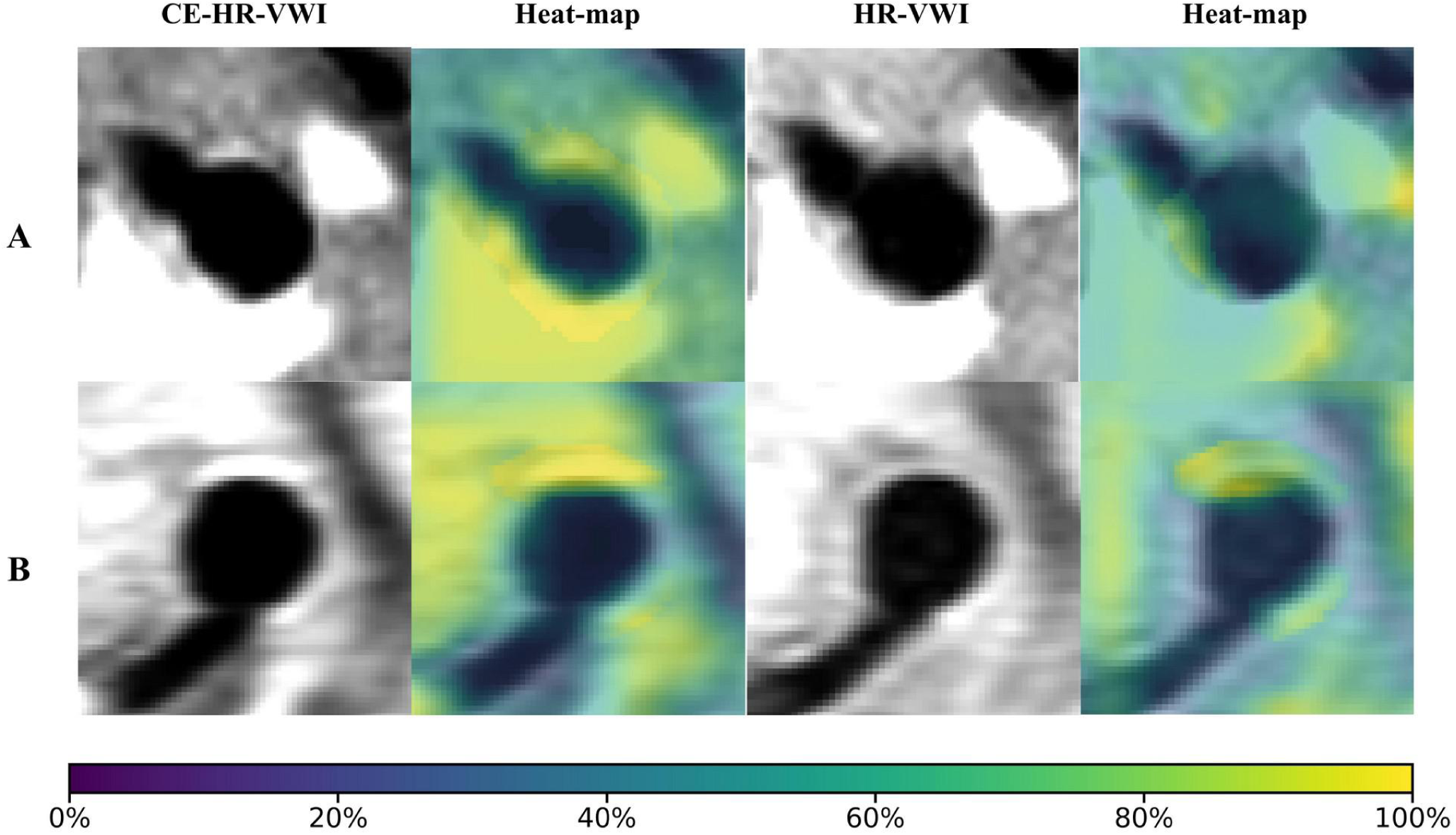
Representative two-dimensional Grad-CAM heatmaps of aneurysms with higher and lower model-predicted probabilities on dual-phase HR-VWI. Two-dimensional Grad-CAM heatmaps are shown on non-enhanced and contrast-enhanced HR-VWI.

## Supplementary Appendix 2. Subset-based ablation analyses for model design

To further evaluate the relative contributions of different model components, subset-based ablation analyses were performed during the preliminary model development stage using the first 200 HR-VWI cases selected chronologically from the internal training cohort. Case selection was based only on enrollment order and was not based on outcome labels, model predictions, or model performance. This subset included 160 negative and 40 positive cases. All ablation models were trained and evaluated for the same binary task: classification of stable versus composite unstable aneurysms. All experiments used a five-fold out-of-fold cross-validation strategy: in each fold, 80% of the samples were used for model training, and predictions were generated for the remaining 20% unseen samples. Predictions from the five folds were then aggregated to calculate overall performance.

Because these analyses were based on a 200-case subset of the training cohort, the results were intended to provide an exploratory assessment of model design rather than independent validation of final model performance. Model comparisons were primarily based on AUC and paired bootstrap AUC comparisons. Average precision, Brier score, and expected calibration error were used as auxiliary metrics to summarize discrimination and probability calibration across ablation models.

### 2.1. Wall-aware feature aggregation ablation analysis

We first evaluated the role of the wall-aware modeling strategy. Wall-aware models extracted features from the local vessel wall region and used the aneurysm wall mask for mask-aware pooling to guide feature aggregation. The corresponding no-wall models did not use mask-aware pooling, while the imaging phase inputs, network branch configuration, and training strategy were otherwise kept unchanged. This ablation analysis was designed to explore whether explicit vessel wall region modeling helped the model use local lesion-related information on HR-VWI.

The wall-aware strategy was associated with higher AUCs in the CE-only, NC-only, and CE+NC fusion settings. In the CE-only setting, the wall-aware stacker achieved an AUC of 0.781, compared with 0.693 for the no-wall stacker. In the NC-only setting, the wall-aware stacker achieved an AUC of 0.780, compared with 0.672 for the no-wall stacker. For the CE+NC fusion model, the four-stream wall-aware stacker achieved a higher AUC in this subset (0.853) than the no-wall fusion model (0.701). The main discrimination and calibration metrics of the ablation models are shown in Supplementary Table S3.

**Supplementary Table S3.** Exploratory performance metrics of ablation models in the 200-case subset.

| Input setting | Aggregation | AUC (95% CI) | Brier score | ECE |
| --- | --- | --- | --- | --- |
| CE-only | No-wall | 0.693 (0.605-0.779) | 0.160 | 0.101 |
| CE-only | Wall-aware | 0.781 (0.696-0.857) | 0.136 | 0.096 |
| NC-only | No-wall | 0.672 (0.576-0.754) | 0.158 | 0.063 |
| NC-only | Wall-aware | 0.780 (0.710-0.850) | 0.154 | 0.096 |
| CE+NC fusion | No-wall | 0.701 (0.613-0.785) | 0.160 | 0.085 |
| CE+NC fusion | Wall-aware | 0.853 (0.790-0.911) | 0.148 | 0.137 |

**Note:** Values are based on aggregated 5-fold out-of-fold predictions in the 200-case internal training subset. ECE = expected calibration error.

### Supplementary Appendix 2.2. Non-enhanced and contrast-enhanced HR-VWI ablation analysis

We further compared the contributions of different imaging phases to model performance, including CE-only, NC-only, and CE+NC fusion models. The CE-only model used only contrast-enhanced HR-VWI images, the NC-only model used only non-contrast-enhanced HR-VWI images, and the CE+NC fusion model integrated both contrast-enhanced and non-contrast-enhanced image information. This experiment was designed to explore whether dual-phase HR-VWI provided complementary predictive information.

Under the wall-aware setting, the CE-only and NC-only models showed similar performance. The CE-only wall-aware stacker achieved an AUC of 0.781, and the NC-only wall-aware stacker achieved an AUC of 0.780, suggesting that both contrast-enhanced and non-contrast-enhanced HR-VWI contained outcome-related discriminative information. The CE+NC fusion model achieved a higher AUC in this subset, with an AUC of 0.853 for the four-stream wall-aware stacker, exceeding those of the CE-only and NC-only models. For average precision, the CE-only, NC-only, and CE+NC fusion models achieved values of 0.500, 0.423, and 0.517, respectively, with the fusion model again showing the highest value.

**Supplementary Table S4.** Exploratory paired bootstrap AUC comparisons for ablation analyses.

| Comparison | AUC A | AUC B | Delta AUC | 95% CI | Bootstrap p |
| --- | --- | --- | --- | --- | --- |
| CE wall-aware vs CE no-wall | 0.781 | 0.693 | +0.088 | -0.020 to 0.187 | 0.110 |
| NC wall-aware vs NC no-wall | 0.780 | 0.672 | +0.108 | -0.002 to 0.224 | 0.056 |
| CE+NC wall-aware vs CE+NC no-wall | 0.853 | 0.701 | +0.153 | 0.052 to 0.254 | <0.001 |
| CE+NC wall-aware vs CE-only wall-aware | 0.853 | 0.781 | +0.073 | 0.009 to 0.138 | 0.022 |
| CE+NC wall-aware vs NC-only wall-aware | 0.853 | 0.780 | +0.073 | 0.013 to 0.134 | 0.017 |

**Note: Model comparisons were conducted as exploratory paired bootstrap AUC comparisons within the 200-case subset. CI = confidence interval. These analyses were used to support model design choices and were not treated as independent validation results.**

Exploratory bootstrap comparisons showed that wall-aware models had higher AUCs than no-wall models in both the CE-only and NC-only settings, with increases of 0.088 and 0.108, respectively. In the CE+NC fusion setting, the wall-aware model had an AUC increase of 0.153 compared with the no-wall model (bootstrap p < 0.001). In addition, the CE+NC wall-aware fusion model showed higher AUCs than both the CE-only wall-aware model and the NC-only wall-aware model (Delta AUC = 0.073 for both comparisons). These exploratory results suggest that CE and NC images both contain discriminative information related to the binary composite instability endpoint and may provide complementary information, supporting the use of a dual-phase HR-VWI fusion strategy in the final model.

